# Frequencies of *TP53* Germline Variations and Identification of Two Novel 3’UTR Variants in a Series of Head and Neck Cancer Cases

**DOI:** 10.1101/2021.09.09.21263314

**Authors:** Sadia Ajaz, Rabbia Muneer, Aisha Siddiqa, Muhammad Ali Memon

## Abstract

TP53 is a tumour suppressor gene. Its inactivation plays a significant role in the molecular pathology of cancers. TP53 germline mutations increase the risk of developing multiple primary cancers. However, the role of alterations in TP53 germline DNA in head and neck cancers (HNCs) is not well-established. HNCs comprise one of the most frequent cancers in South Asia. The present discovery study reports the investigation of germline variations in the TP53 gene in a cohort of 30 HNC patients from Karachi, Pakistan. Blood samples were collected and genomic DNA was extracted from white blood cells. TP53 has 11 exons, where exon 1 is not transcribed. After quality control of DNA, amplification of seven selected exons along with their splice sites, two intronic regions (introns 2-3 and 3-4), and 3’UTR were carried out. Sanger sequencing was carried out in order to identify germline variations. Comparison with wild type sequence revealed rs1642785 G>C (intron 2-3) variation in 63.2%, PIN3 duplication (rs17878362) in intron 3-4 in 94.7%, and rs1042522 G>C in exon 4 (p.R72P) in 66.6% of the cases. In 3’UTR, 13.4% of the analyzed cases carried either one of two variants, i.e., 17:7669567_8delCA or 17:7669560C>G. The latter variations are reported for the first time in literature. In conclusion, we report three highly frequent germline variations and two newly discovered variations in 3’UTR of TP53 germline DNA in HNC patients from Pakistan. These results shall contribute to delineating the genetic component of HNCs with potential translational implications.

## Introduction

The locus of *TP53*, a tumor suppressor gene, is 17p13.1. It covers ∼ 25.76 kb of DNA. The transcript length is 2,512 bp. It consists of 11 exons, where 10 exons constitute the coding sequence of p53. The protein is made up of 393 amino acids (Ensembl, 2021). p53 is known as the guardian of the genome (Bergh, 1999). It mainly functions as a tumor suppressor and a transcription factor. It plays a key role in the signaling pathways coordinating cellular response to stress and damage. It is a major regulator of apoptosis, autophagy, angiogenesis, differentiation, DNA repair, ferroptosis, metabolism, proliferation and senescence (Monti *et al*., 2020).

As shown in Figure 1, p53 consists of distinct domains. The <u>A</u>mino <u>T</u>erminal domain (AT) is encoded partly by exon 2, where translation initiation codon is present. The remaining component of AT domain is encoded by exon 3. This domain includes two functionally significant trans<u>a</u>ctivation <u>d</u>omains (TAD1 and TAD2) (Raj and Attardi, 2017). The <u>P</u>roline <u>R</u>ich <u>D</u>omain (PRD) is encoded by exon 4. A folded DNA <u>B</u>inding <u>D</u>omain (DBD), which plays a critical role in the transcription activation function of p53, is encoded by exons 4 - 8. A short region of <u>N</u>uclear <u>L</u>ocalization <u>S</u>ignal (NLS) is also encoded in exon 8. <u>C</u>-<u>T</u>erminal regulatory <u>D</u>omain (CTD) containing tetramerization or <u>O</u>ligomerization <u>D</u>omain (OD) and Basic Repression (BR) domain, is encoded by exons 9-11. A <u>N</u>uclear <u>E</u>xport <u>S</u>ignal (NES) is also present in CTD. Genetic alterations in exons, introns, and 5’, 3’ untranslated regions (UTRs) lead to highly specific aberrations in p53 function.

**Figure 1.**
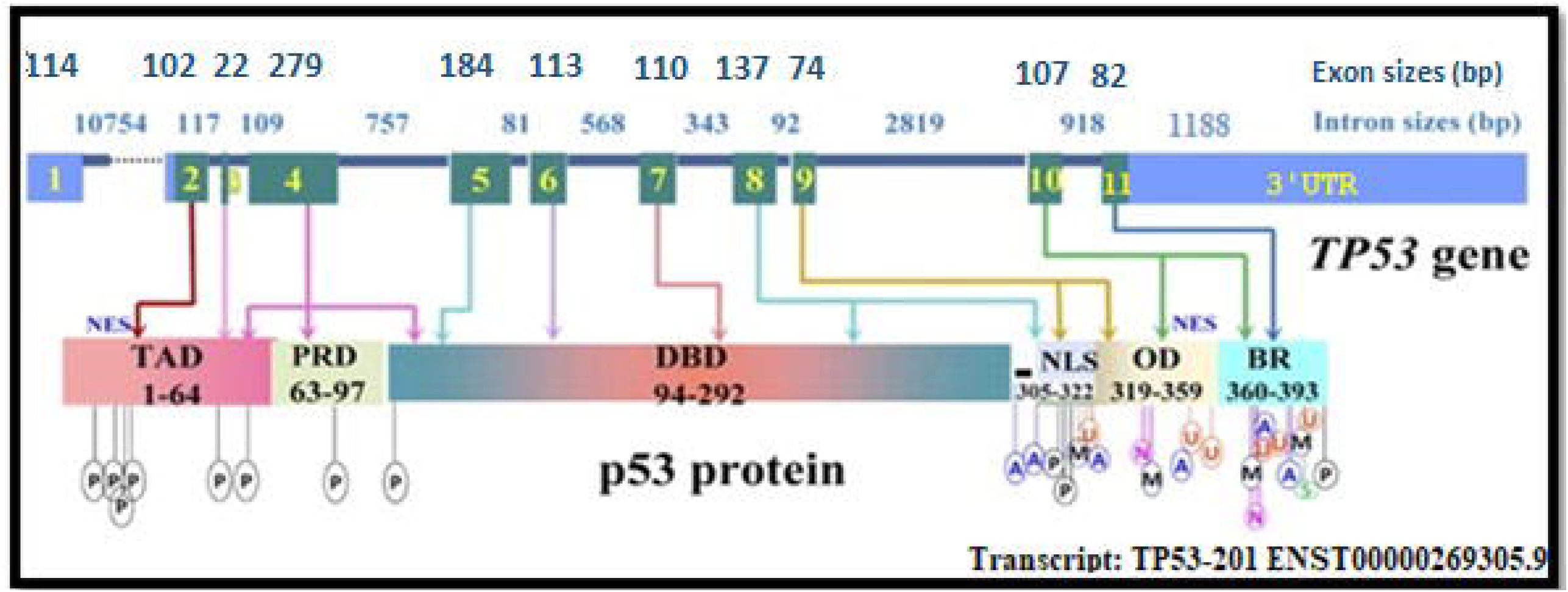
*TP53* gene and p53 protein functional domains. NES are present on TAD and OD. NES: Nucleus Export Signal. TAD: Transactivation Domain. PRD: Proline Rich Domain. DBD: DNA Binding Domain. NLS: Nuclear Localization Signal. OD: Oligomerization Domain. BR: Basic Repression Domain. Posttranslational modification of p53 includes Phosphorylation (P), Ubiquitination (U), Acetylation (A), Sumoylation (S), Methylation (M), Neddylation (N). [adapted from (Xu-Monette *et al*., 2012)].

*TP53* is the most frequently mutated gene in cancers. It is well known for germline and somatic variations in inherited and sporadic tumors, respectively (Bouaoun *et al*., 2016). The inherited conditions associated with germline mutations in *TP53* include Li-Fraumeni syndrome and Osteosarcoma (OMIM 191170). Previously, 0.31% of head and neck cancers (HNCs) have been associated with germline mutations in *TP53* (IARC, 2019). Among HNC patients, *TP53* mutation is a common somatic alteration (Zhou, Liu, and Myers, 2016).. However, little literature is available for the frequency of germline *TP53* variations in the HNCs. The present study addresses this gap. Such variations may modify the cancer-associated parameters such as etiology, pathology, and response to treatment. It investigates germline DNA variations in functionally significant exons, the reported highly polymorphic intronic regions, and 3’UTR of *TP53* gene in HNC patients.

## Methodology

### Ethics Statement

The study was carried out in accordance with Declaration of Helsinki (WMA, 2018). Details of ethical approval for the study and participants’ informed consent have been published earlier (Ajaz *et al*., 2021).

### Blood sampling and DNA extraction

In this discovery study, 8-10 ml of peripheral blood samples were collected in ACD coated vacutainers (BD^®^, Inc.) from 30 HNC patients. DNA extraction from white blood cells was carried out by using standard phenol/chloroform method (Russell and Sambrook, 2001). Final DNA precipitation was carried out in an equal volume of isopropanol with 1/10th volume of 10M ammonium acetate. The precipitates were washed with 70% ethanol and re-suspended in 10mM Tris-EDTA (TE) buffer. DNA purity and concentration were determined spectrophotometrically and sample stocks were stored at - 20ºC. The 40ng/μL dilutions were prepared for subsequent PCRs.

### Identification of germ-line variations

The amplification of targeted sequence was done through PCR. The details are given below:

#### PCRs for *TP53* germline variations

PCRs were carried out with reagents from Thermo Fisher Scientific, Inc. Touch down PCRs were carried out in a thermal cycler (Kyratec^®^). Primer sequences (IARC, 2010) for the selected exons and introns are listed in Table 1. Total volume for each PCR was 25μL. PCR conditions for each exon are given below:

**Table 1.**
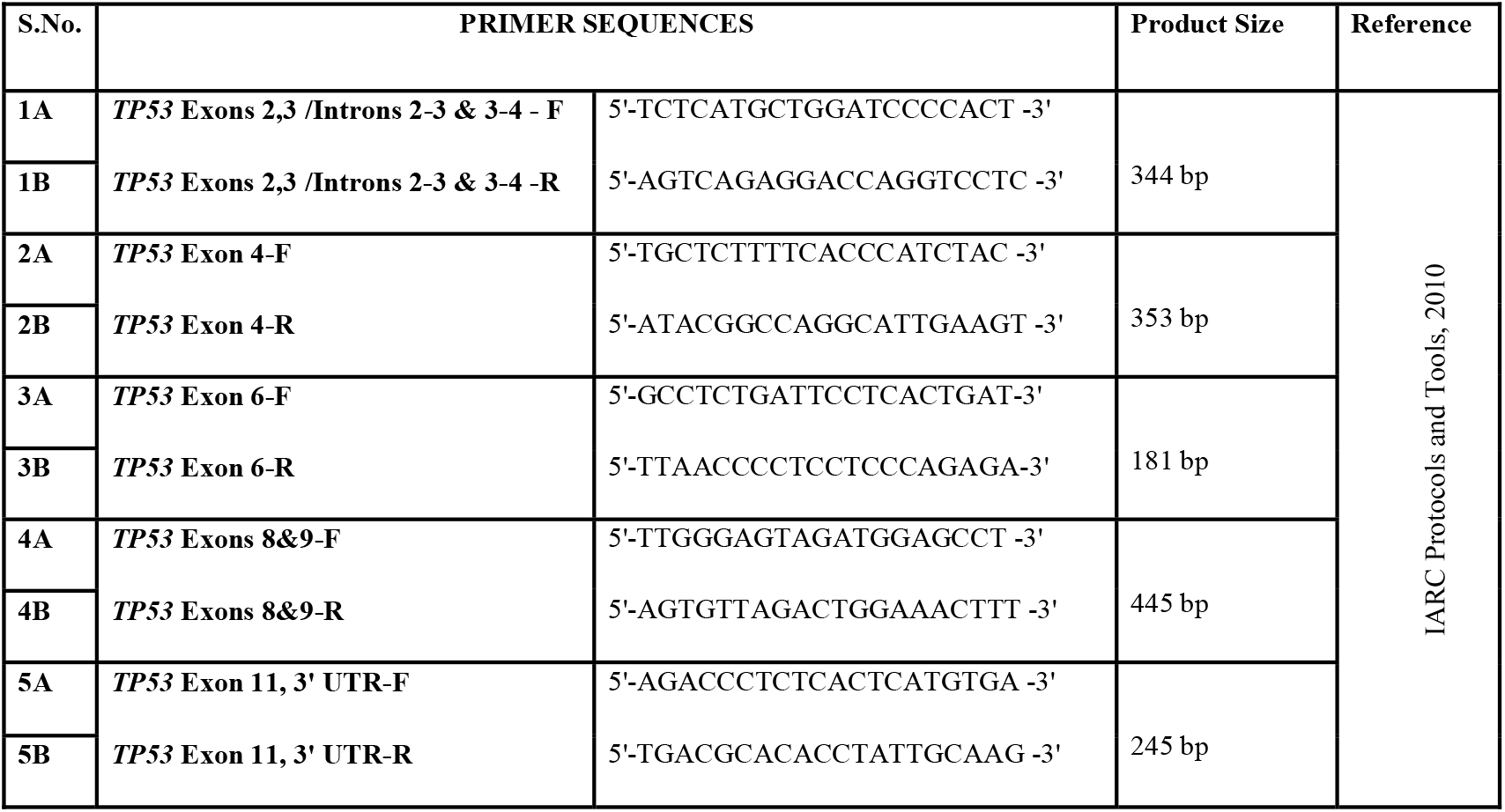
Primer sequences for the amplification of *TP53* exons.

i. *<u>TP53</u>* <u>Exons 2 & 3; Introns 2-3 & 3-4</u>: 1X PCR Buffer with (NH_4_)_2_SO_4_, 1.1mM MgCl_2,_ 0.2mM dNTPs, 0.5U of *Taq polymerase*, 0.4μM *TP53* exons 2,3/introns 2-3 and 3-4 forward, reverse primers, and 80ng total genomic DNA.
ii. *<u>TP53</u>* <u>Exon 4</u>: 1X PCR Buffer with (NH_4_)_2_SO_4_, 1.5mM MgCl_2,_ 0.2mM dNTPs, 0.5U of *Taq polymerase*, 0.4μM *TP53* exon 4 forward and reverse primers, and 100ng total genomic DNA.
iii. *<u>TP53</u>* <u>Exon 6</u>: 1X PCR Buffer with (NH_4_)_2_SO_4_, 1.5mM MgCl_2,_ 0.2mM dNTPs, 0.5U of *Taq polymerase*, 0.4μM *TP53* exon 4 forward and reverse primers, and 100ng total genomic DNA.
iv. *<u>TP53</u>* <u>Exons 8 & 9</u>: 1X PCR Buffer with (NH_4_)_2_SO_4_, 0.87mM MgCl_2,_ 0.2mM dNTPs, 0.5U of *Taq polymerase*, 0.4μM *TP53* exons 8 & 9, forward and reverse primers, and 80ng total genomic DNA.
v. *<u>TP53</u>* <u>Exon 11 & 3’UTR</u>: 1X PCR Buffer with (NH_4_)_2_SO_4_, 1.15mM MgCl_2,_ 0.2mM dNTPs, 0.5U of *Taq polymerase*, 0.4μM *TP53* exon 11 & 3’UTR forward, reverse primers, and 100ng total genomic DNA.

PCR thermal cycling conditions for all exons were the same. Touch down PCRs were carried out with initial denaturation at 94ºC for 2 min followed by 20 cycles (94 ºC for 30 sec. 63 ºC for 45 sec. and 72 ºC for 1 min), 30 cycles (94 ºC for 30 sec. 60 ºC for 45 sec. and 72 ºC for 1 min) and final elongation at 72 ºC for 10 min. 5μL of the amplified products in 6X DNA gel loading dye were run on 2.5% agarose gels with ethidium bromide along with 50bp ladder. The amplicons were observed in Azure gel imaging system under UV light.

#### DNA Sequencing and Identification of Variants

After quality control for PCR amplification, the amplicons were purified with isopropanol. Sanger sequencing for selected exons, introns, and 3’UTR was carried out commercially. The variants were identified by comparing the sequences for HNC patients with reference *TP53* sequence (HGNC:11998)

## Results

### Demographic and Clinico-pathological Characteristics of HNC Patients

The study analyzed DNA samples from 30 HNC patients. In this pilot study, 53.3% (16/30) patients were male, while 46.7% (14/30) patients were female.

Age-wise comparison showed that 69% patients were above the age of 40 years and 31% patients were less than or equal to the age of 40 years. In addition, 65% of the analyzed patients reported addiction to the known risk factors for HNCs, i.e., betel leaf, betel nut, tobacco, and or smokeless tobacco, while 35% reported no such addiction.

Among HNC cases, 58.6% had oral cavity cancer, followed by 17.2% cases each of oropharyngeal and nasopharyngeal carcinoma. Squamous cell carcinoma was the most frequently reported histological sub-type in this studied cohort (83.2%).

Family history assessment showed that out of 30 patients, only one patient had positive history of familial HNC.

### Identification of Polymorphisms and Variants in *TP53* Gene

The schematic gene structure of *TP53* with identified variants, marked investigated and un-investigated regions is presented in Figure 2. The details of identified germline variants are given below:

**Figure 2.**
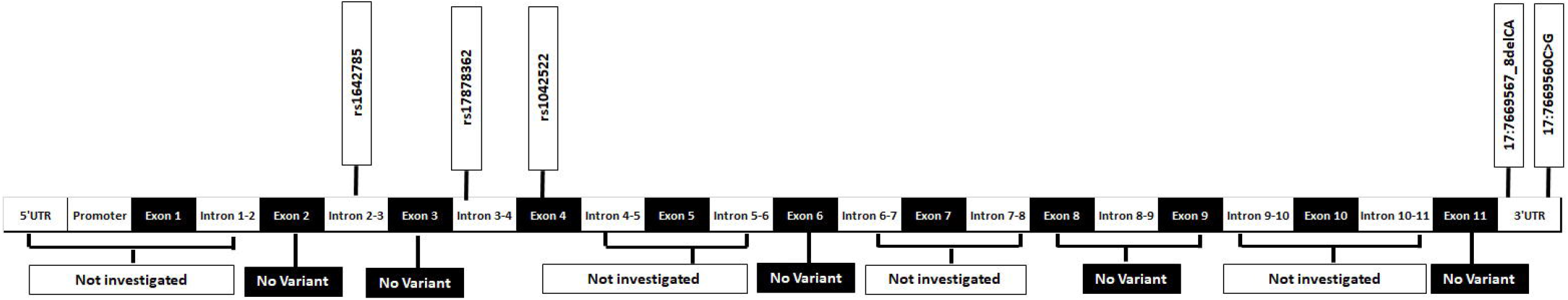
Schematic diagram of *TP53* gene structure.

### rs1642785 G>C (Intron 2-3) variation

This polymorphism was identified in 63.2% of the analyzed cases (heterozygous and homozygous carriers of the variant). Representative relevant component of the electropherogram is shown in Figure 3.

**Figure 3:**
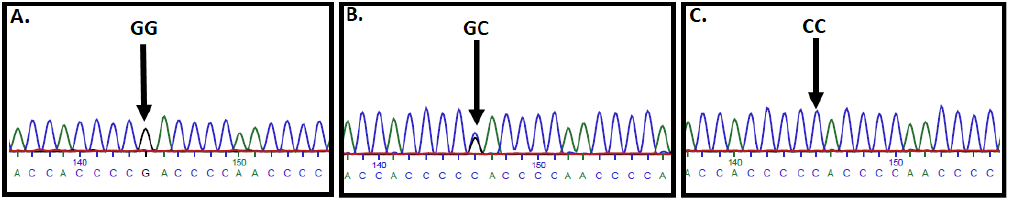
Sanger sequencing displaying *TP53* rs1642785 polymorphism indicated by arrow. (A.) Wild type [GG], (B.) Heterozygous [GC], (C.) Variant [CC].

The frequencies of homozygous G, GC, and homozygous C for rs1642785 were 33.3%, 47.6%, and 9.5%, respectively.

### rs17878362 [Polymorphism in Intron 3 (PIN3) Duplication]

In intron 3, PIN3, a 16bp duplication (rs17878362) was detected in 94.7% of the HNC cases (Figure 4). The frequency of homozygous carriers of PIN3 duplications was 89.5%, whereas the frequency of heterozygous PIN3 duplication carriers was 5.3%.

**Figure 4:**
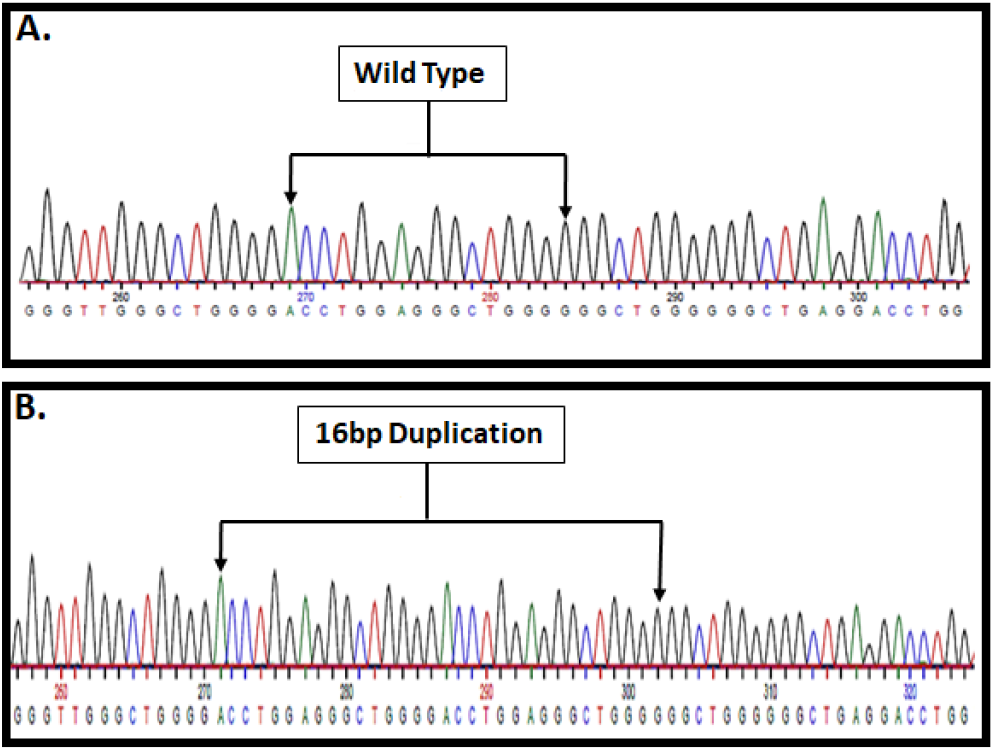
Sanger sequencing of *TP53* distinguishing between wild-type sequence (A.) and polymorphism in intron 3 (PIN3), a 16 bp duplication (5′-ACCTGGAGGGCTGGGG-3′) (B.).

### rs1042522 G>C Polymorphism in Exon 4 (p.R72P)

rs1042522 in exon 4, which substitutes proline for arginine at codon 72, was identified in 66.6% of the cases (Figure 5).

**Figure 5:**
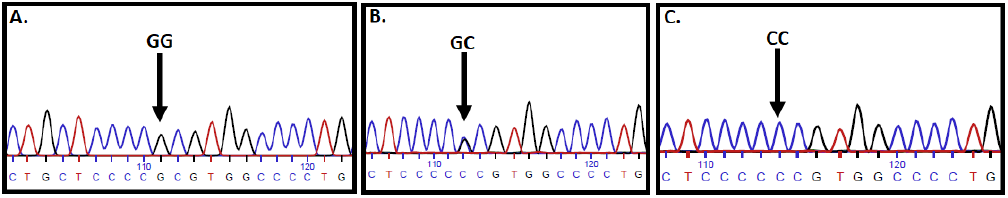
Sanger sequencing displaying *TP53* rs1042522 polymorphism indicated by arrow. (A.) Wild type [GG], (B.) Heterozygous [GC], (C.) Variant [CC] sequences.

The frequencies of homozygous G, GC, and homozygous C for rs1042522 were 33.3%, 60%, and 6.6% respectively.

### 3’UTR 17:7669567_8delCA and 17:7669560C>G

Among 30 cases, a new variant 17:7669567_8delCA was detected in 3.3% of cases. Another variant in this genomic region, which is reported for the first time in literature, 17:7669560C>G, was observed in 10% of the cases (Figure 6)

**Figure 6:**
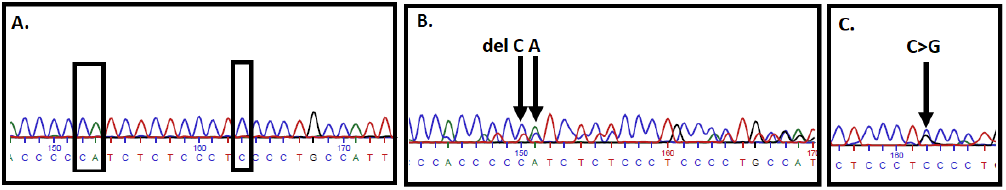
Representative Sanger sequencing displaying (A.) Wild type sequence in 3’UTR, (B.) del CA at chromosome 17:7669567_8 and (C.) C>G at chromosome 17:7669560.

## Discussion

Tumour progression depends on mutant p53, which promotes cancer cell proliferation and growth. It may also lead to poor therapeutic response (Sabapathy and Lane, 2018).

AT domain, DBD, and OD are the most common sites of mutations (Liu *et al*., 2019). The present study reports only one germline variation in the exonic region. This variant is located in the genomic region encoding PRD. The remaining four variations were identified in either introns or 3’UTR.

The functions of discrete domains in p53 and the probable consequences of the identified variations in the present study are discussed below:

### Amino Terminal Domain [Identified Polymorphisms: rs1642785 G>C and rs17878362: PIN3]

*TP53* exons 2 and 3 encode for AT domain of p53. The translation initiation codon ATG for p53 is present in exon 2 (translation of p47 - an amino terminal truncated 47 kDa protein - starts from another start codon which is located in *TP53* exon 4) (Phang *et al*., 2015). The AT domain consists of two short amphipathic motifs, termed as Transactivation Domains, “TAD1” and “TAD2”, which contain 1–40 and 41–61amino acid residues, respectively (Krois, Dyson, and Wright, 2018; Sabapathy and Lane, 2018).

TAD forms binding sites for various regulatory proteins. The TAD motifs fold into ordered helical structure upon binding to regulatory partners (Krois *et al*., 2016). These interactions are important for p53 function as well as negative regulation, proteasomal degradation and stabilization, and p53 targeted genes activation. As the cellular stress level increases, phosphorylation of TAD also increases as shown in Figure 1 (Krois *et al*., 2018).

Deletion and rearrangement in TAD domain results in loss of p53 protein expression. (Phang *et al*., 2015).

The polymorphisms identified in the present study (rs1642785 and rs17878362) are reported to be in linkage disequilibrium (Perriaud *et al*., 2014). They are known to effect mRNA spicing. The presence of rs1642785-C allele leads to lower transcript stability. This allele, irrespective of rs17878362-PIN3 variant, leads to lower p53 pre-mRNA, total TP53, fully spliced p53 transcript (FSp53) and the N-terminally truncated Δ40p53 isoform (p53I2) levels. On the other hand, the combination of the wild-type variants for both rs1642785 and rs17878362 leads to the highest levels of both FSp53 and p53I2. The homozygous C genotype for rs1642785 has been shown to be associated with the risk for chronic lymphocytic leukemia (CLL) (Bilous *et al*., 2017).

PIN3 is a 16 bp (5′-ACCTGGAGGGCTGGGG-3′) duplication (rs17878362). The intron 3-4 is relatively small in size i.e. 93 bp. The 16 bp insertion in intron 3-4 may lead to 17% increase in the length of this intron. This insertion is known to alter gene expression and protein function (Winiecka-Klimek *et al*., 2014). PIN3 duplication has shown association with breast and colorectal cancer risk (Sagne *et al*., 2013, Diakite *et al*., 2020).

In Pakistani population, the frequencies of rs1642785 and rs17878362 variants in germline DNA of HNC patients were 63.2% and 94.7%, respectively. As already discussed these *TP53* polymorphisms may contribute in cancer progression via either the transcript stability or alternative splicing.

PIN3 heterozygotes are responsible for *TP53* haploinsufficieny (Winiecka-Klimek *et al*., 2014). Haploinsufficiency leads to preferential transcription of the variant allele. Thus, the expression of wild-type allele is suppressed. In the present study only one patient was identified with heterozygosity for PIN3.

PIN3 heterozygosity frequencies have been reported previously in healthy populations. In a study from South-Asia comprising 80 females, it was 31% (Guleria *et al*., 2012), 23% in South American population (Paskulin *et al*., 2012) and 21% in an European population (Gemignani *et al*., 2004). In the present study, only one case was identified with PIN3 heterozygosity (5.2%). The majority of patients (89.5%) were homozygous for PIN3 variant. Therefore, the role of PIN3 haploinsufficiency in HNC cases remains uncertain.

### Proline Rich Domain [Identified Polymorphism: rs1042522 G>C (p.R72P)]

Proline rich region (PRD) is encoded by exons 3 and 4 of *TP53*. PRD connects TAD to DBD. It carries 12 proline amino acids. It promotes protein–protein interactions in signal transduction. With rs104252 variant, although other domains of p53 remain conserved; however, PRD undergoes functional alteration. Conserved PRD plays important role in apoptosis and growth suppression (Chillemi *et al*., 2017).

rs1042522 (p.R72P) effects p53 structure. It presents in the PRD of p53. This domain regulates apoptosis and growth suppression of cell (Doffe *et al*., 2021; Almutairi *et al*., 2021). The SNP results in the missense substitution of proline (Pro) by arginine (Arg) (dbSNP, 2021).

An earlier study found that Arg/Arg homozygotes induce apoptosis more efficiently as compared to Pro/Pro homozygotes through the arrest of cell cycle in G1 phase (Pim and Banks, 2004), however, this observation is not supported by a recent study where wild-type p53 was indistinguishable from the variant forms in its apoptotic function (Doffe *et al*., 2021).

The association of codon 72 polymorphism with breast, colorectal, ovarian, and endometrial cancers remains controversial (Dahabreh *et al*., 2013). p53 protein with Arg at codon 72 is more likely to be degraded by E6 protein of HPV11, HPV16, HPV18, and thus facilitates the oncogenic consequence of HPV (Storey *et al*., 1998). Interestingly, in the absence of HPV, Arg allele appears to provide protection against HNCs (Cortezzi *et al*., 2004).

In the present study, we report the G-variant (Arg-variant) frequency of 66.6% in HNC cases for p.R72P. In comparison with South Asian (SAS) population in 1000G project, which reports frequencies of 24.3%, 49.7%, and 26%, for homozygous G, GC, and homozygous C genotypes, the present study reports these frequencies as 33.3%, 60%, and 6.6%, respectively. Similarly, the frequencies of G-allele and C-allele are reported respectively as 49% and 51% in 1000G project, while in the present study, the respective frequencies were 63.3% and 36.7%. These comparisons indicate that Arg substitution at codon 72 may be a risk factor for HNC in South-Asian populations. However, this observation is in contrast to the previous studies, which imply that Pro72 in p53 may be associated with increased risk to cancers (Cao *et al*., 2020, Lotfi Garavand *et al*., 2020, Pinheiro *et al*., 2015, Cortezzi *et al*., 2004). This contradiction may be partially explained by other genetic and/or environmental factors, which have not been considered in the present discovery study.

### DNA Binding Domain [Investigated Exons: 6 and 8]

Albeit the sample size was small, we did not find any germline variant in the exons 6 and 8 which partially encode for DBD. The reported *TP53* mutations, mainly from Caucasians, most frequenctly occur in the DBD. This domain contains 540 nucleotides with numerous recurring hotspot mutations including frameshift, nonsense, silent, and missense mutations while the missense mutation is more frequent in DBD. DBD mutant p53 is relatively a stable protein with the loss of WT functions. In addition to loss of function (LOF), DBD mutations also gain additional oncogenic properties. Exon 8 was identified as the most frequently affected site in DBD (Liu *et al*., 2019). We do not report any germline mutation/variant in the discovery analysis for the DNA of this exon.

DBD is encoded by *TP53* exons 5 to 8 (Kadia *et al*., 2016). It consists of 200 amino acids. DBD regulates the transactivation and DNA binding functions of p53, other functions include DNA binding ability, apoptosis, cell cycle arrest, and RAS-induced senescence as well as tumor suppressor activity. Thus, it plays crucial role in tumor suppression (Hanel *et al*., 2013).

Almost 80% of all mutations reported in *TP53* mutations occur within DBD (Bouaoun *et al*., 2016). p53 DBD mutants have lost the potential to trans-activate its target genes (Lee *et al*., 2012). Therefore, mutations in the DBD results in loss of p53 function.

*TP53* germline mutations in the DBD predispose to spontaneous cancer development typically in individuals with Li-Fraumeni syndrome (Hanel *et al*., 2013).

Mutant DBD produces “Dominant Negative” effect. If the mutant p53 interferes the wild type (WT) protein activities in the heterozygous state and inactivates them is known as Dominant Negative effect. This dominant negative effect occurs at the early stages of cancer. Mutation in one allele reduces the overall function of p53 and results in haploinsufficieny (Sabapathy, 2015).

In the later stages of cancer progression, after the Loss of Heterozygosity (LOH), mutant p53 is unable to recognize its target sequence resulting in the loss of WT functions. Some p53 mutant have acquired novel gain-of-function (GOF) activities, which in turn increases cancer progression, metastasis and genomic instability.

The gain-of-function or oncogenic functions of mutant p53 includes, angiogenesis, chemo-resistance, invasion, migration, mitogenic defects, proliferation, scattering, stem cell expansion, survival, and tissue remodeling. These wide variety of responses showed that mutant p53 can function through several different pathways (Muller and Vousden, 2013). Thus, p53 is also called “contrived” oncogene (Sabapathy, 2015).

### Carboxyl Terminal Domain [Investigated Exons: 9 and 11)

*TP53* exons 9 to 11 encodes for the carboxyl terminal domains of p53. It consist of an Oligomerization Domain (OD) and Basic Repression domain (BR) (Joruiz and Bourdon, 2016).

#### Oligomerization Domain (OD)

The Oligomerization Domain (OD) is encoded by *TP53* exons 9 to 10. It comprises of 325–356 amino acids of p53. These amino acids are important for the p53 tetramerization, which is the critical step for tumor suppressor activity (Sabapathy and Lane, 2018). OD undergoes alternative splicing and posttranslational modifications (Kiraz *et al*., 2016).

Various *TP53* germline mutations have been identified within the OD that has an association with Li-Fraumeni syndrome. The connection between OD mutations and LFS shows the importance of OD in maintaining the cellular homeostasis (Fischer *et al*., 2016).

Consequently, mutation in the OD acquired p53 LOF. These OD mutant proteins have lost the potential to function as tumor suppressors or trans-activate the target genes (Fischer *et al*., 2016). Mutations that disturb p53 tetramerization also abrogate DNA binding, hence reducing the target gene activation (DiGiammarino *et al*., 2002). Conventional therapies may not be effective for the patients with OD mutations. These patients rely on p53-dependent cytostatic or cytotoxic effect for the eradication of cancer cells (Sabapathy and Lane, 2018). Rare OD mutations are also present in sporadic cancers (Bouaoun *et al*., 2016). OD plays important role in tetramerization of p53. OD mutations often act as LOF mutations. Patients with OD mutations are less sensitive to therapies. These patients are mostly dependent on p53-mediated cytotoxicity (Sabapathy and Lane, 2018).

We do not report any germline variant in exon 9 in the present series of HNC cases.

### Basic Repression domain

*TP53* exons 10 and 11 encodes the Basic Repression (BR) domain. It is rich in basic amino acid lysine. It undergoes several posttranslational modifications including, acetylation, phosphorylation, sumoylation, methylation, ubiquitinylation, neddylation, etc. It regulates p53 stability and activity (Joruiz and Bourdon, 2016).

Mutations in *TP53* downregulates apoptosis that leads to unstable genome and results in loss of tumor suppression activity (Muller and Vousden, 2013). These alterations also induce gain of function mutations that facilitate tumor progression as well as metastasis (Liu *et al*., 2019).

The present study did not identify any germline variant in *TP53* exon 11.

In 3’UTR, novel deletion delCA and G>C variants were identified, which might adversely affect the protein translation.

All variations require validation by reverse sequencing.

In the literature PIN3 and codon 72 Pro/Arg are the most commonly reported polymorphisms. The position and the type of mutation in *TP53* is important as it has differential effects on prognosis (Canale *et al*., 2017). The literature shows that different p53 mutant forms have several important functional differences.

## Conclusion

In conclusion, we report three highly frequent germline variations and a newly discovered variation in 3’UTR in *TP53* germline DNA of HNC patients from Pakistan. Further functional analysis is recommended to assess the effect of these variations in cancer etiology and pathology. Due to the high incidence and prevalence of HNCs in this region, this trend needs to be explored further

## Supporting information

STROBE CHECKLIST

## Data Availability

Data is available on request.

